# Development of an AI-enabled predictive model to identify the ‘sick child’ at a pediatric telemedicine and medication delivery service in Haiti

**DOI:** 10.1101/2025.06.27.25330413

**Authors:** Ben J. Brintz, Molly B. Klarman, Youseline Cajusma, Lerby Exantus, Jude Ronald Beausejour, Katelyn E. Flaherty, Valery Madsen Beau de Rochars, Chantale Baril, Eric J. Nelson

## Abstract

**Background:** One of the most difficult challenges in pediatric telemedicine is to accurately discriminate between the ‘sick’ and ‘not sick’ child, especially in resource-limited settings. Models that flag potentially ‘sick’ cases for additional safety checks represent an opportunity for telemedicine to reach its potential. However, there are critical knowledge gaps on how to develop such models and integrate them into electronic clinical decision support (eCDS) tools.

**Methods:** To address this challenge, we developed a study design that utilized data from paired virtual and in-person exams at a telemedicine and medication delivery service (TMDS) in Haiti. Providers were allowed to mark respondent data as potentially unreliable. Artificial intelligence /machine learning (XGBoost) was applied to analyze paired data from participants across three consecutive implementation studies. Model derivation focused on identifying ‘sick’ patients (not-mild) and those requiring escalation. An ensemble method, based on gradient boosted decision-trees, was used given the limited sample size. The area under the receiver operating characteristic curve (AUC) was the primary outcome measure.

**Results:** A total of 683 paired records were available for this secondary analysis from 2225 participants enrolled. The median age was 15 months and 47% were female. For prediction of a ‘sick’ child, we found an AUC of 0.82 (95% CI 0.78-0.86) after 5-fold cross validation; calibration slope and intercept were 1.31 (95%CI:1.09-1.53) and 0.04 (95%CI:-0.14-0.23), respectively. For prediction of escalation, we found an AUC of 0.78 (95%CI:0.74-0.81); calibration slope and intercept were 0.63 (95%CI:0.52-0.74) and 0.05 (95%CI:0.52-0.74), respectively. Accounting for data marked as potentially unreliable had mixed effects.

**Interpretation:** These methods and findings offer an innovative and important proof-of-concept to improve pediatric telemedicine. The models require external validation prior to eCDS integration and deployment. Once validated, the models are designed to provide a critical safety check for experienced providers and digitally convey expertise to new providers.

**Funding:** National Institutes of Health (USA) grants to EJN (R21TW012332; DP5OD019893), internal funding at UF (Children’s Miracle Network), and private donations.

**RESEARCH IN CONTEXT:** *Evidence before this study:* We conducted two Pubmed searches for reports published in all languages. The first search terms were (telemedicine) AND (artificial intelligence OR machine learning) AND (pediatrics OR paediatrics). The primary search criteria identified 153 publications and reviews were excluded leaving 101 papers all published after 1998. Enumerated results by named subspecialty were neurology (n=2), ophthalmology (n=13), otology (n=3), endocrinology (n=11), cardiology (n=6), pulmonology (n=3), gastroenterology(n=1), dermatology (n=1) and surgery (n=22). Ten of the publications focused on global health or low-middle income countries (LMIC) populations. The second search terms were ((Telemedicine) AND (delivery OR paramedicine) AND (pediatrics OR paediatrics)) AND (global health OR LMIC) which generated 99 publications and 76 papers remained after reviews were excluded, all published after 2015. After manual evaluation of the results from both searches, no publications were identified that fully met the scope of this paper. Examples of telemedicine research for concordance with paired exams does exist ^1,2^.

*Added value of this study:* To the best of our knowledge, this is the first study that investigated pediatric disease severity prediction using virtual and in-person exams in the context of telemedicine -- for either high or low resourced settings. Therefore, the added value of this study is an innovative and important proof-of-concept to improve telemedicine research and clinical practice beyond the scope of global health.

*Implications of all the available evidence:* Inside the field of global health, there is a need to develop evidence-based approaches to extend care early to pediatric patients who may be isolated by poverty, geography or unrest. This must be done safely and this paper offers an approach to develop and incorporate disease severity prediction models into eCDS tools. In addition, these tools may serve as a welcomed safety check for experienced providers and a method to digitally convey expertise to new providers as these services scale.

## INTRODUCTION

One of the most important and difficult challenges in pediatrics is to correctly discriminate between the ‘sick’ and ‘not sick’ child. This challenge arises in part because children have a great capacity to maintain normal physiologic function until a tipping point is reached when children can rapidly decompensate. This ability to mask an impending crisis is one reason why pediatric training requires mastering how to detect the subtle early patterns of the ‘sick’ child. Achieving this level of clinical acumen takes time and represents one seemingly timeless barrier to providing high-quality pre-emergency pediatric care at scale.

Telemedicine represents a promising method to improve access to pediatric care, especially for low-resource settings. However, accurately identifying the ‘sick’ child in a telemedicine environment is difficult and there is a lack of pediatric telemedicine guidelines. The World Health Organization’s (WHO) integrated management of childhood illnesses (IMCI) guidelines were primarily developed for low-resource settings and are not adapted for telemedicine^3^. The American Academy of Pediatrics (AAP) developed telemedicine guidelines, however they were designed for high-resource settings^4^. One additional limitation with the AAP guidelines is that their threshold for hospital (emergency) referral is low which poses the risk of incurring unnecessary clinical and financial costs. Essentially, the AAP guidelines operate on a red and green stoplight workflow that lacks a yellow option which is a key component to cost-effectiveness^5^.

To address these complex challenges, we launched the Improving Access to Nighttime Care and Treatment (INACT) studies in Haiti^5–9^. We developed a pediatric telemedicine guideline derived from the WHO IMCI guidelines^3^. The guidelines were implemented within a telemedicine and medication delivery service (TMDS) called MotoMeds. The guideline was evaluated for safety by comparing paired virtual and in-person exams. The TMDS was subsequently configured for scaling by triaging cases with higher clinical uncertainty to escalated care that might involve hospital referral (red) or an in-person exam at the household with medication delivery (mostly yellow)^7,8^. Cases with lower clinical uncertainty (mostly green) received medication delivery alone. Analysis revealed a risk that some patients might not be identified as ‘sick’ during virtual exams, despite adherence to the clinical guidelines^6,8,9^. Contributing factors included parent interpretation of provider questions, rigidity of branching logic within the guidelines, and non-clinical situational events. These factors are common across pediatric telemedicine services in both high and low resource settings. They collectively represent a vulnerability that in select situations parents might falsely be reassured that a child is ‘not sick’ based on a virtual exam alone.

We hypothesized that predictive models might address this problem by flagging cases that need added consideration. When integrated into electronic clinical decision support (eCDS) tools, these models could serve as a safety check for experienced providers and digitally convey expertise to new providers. To test this hypothesis, we used paired virtual and in-person exams from the Haitian INACT study data to derive and internally validate predictive models to identify cases that require added consideration. Herein, we demonstrate proofs of concept that data from virtual exams can be leveraged to identify ‘sick’ children in a manner that guidelines might miss.

## METHODS

### Ethics statement

The INACT studies were reviewed and approved by the University of Florida Institutional Review Board (IRB202002693; IRB201802920, IRB202201220) and the Comité National de Bioéthique (National Bioethics Committee of Haiti; 2021-11; Ref1819-51; Ref1101-41).

### Study design

This study represents a secondary analysis of pooled data from three prior prospective cohort studies: INACT2-H established the TMDS model and focused on safety, feasibility and evaluation of the clinical guidelines; all non-severe cases had paired virtual and in-person exams^7,10^. INACT3-H positioned the TMDS for scale by reserving in-person exams for cases with higher clinical uncertainty, which mainly included moderate (yellow) cases^11^. Continuing this scalable approach, INACT4-H is an ongoing interrupted time-series study of a digitization of the TMDS clinical guidelines; analysis draws on data from the pre-intervention period.

### Study population and setting

INACT2-H was conducted in Gressier, Haiti. INACT3-H and INACT 4-H were conducted in both Gressier and Les Cayes, Haiti. The studies were implemented consecutively without interruption. INACT2-H began in September 2019 and concluded in January 2021, followed by INACT3-H, which concluded in September 2022 and the INACT4-H study is ongoing. Gressier has a population of 36,400 and is characterized by rural mountainous regions and more densely populated coastal communities ^12^. Les Cayes is approximately 175km west of Gressier. It has a population of 152,000 people ^12^, and over half live in the urban center. All study periods were marked by political and societal instability with detrimental effects on health and healthcare ^13,14^.

### Participant recruitment

The study populations were informed of the TMDS through radio and print advertisements, plus announcements at schools, clinics and markets.

### Participant inclusion criteria and consent process

For all studies, children were eligible to participate if their parent contacted the TMDS regarding their child aged ≤10 years during operational hours (6 PM-5 AM). Written consent was obtained from parents/guardians, and assent from children ≥7 years, who received an in-person exam. Parents/guardians whose only contact was a virtual encounter were read a waiver of documentation of consent over the phone.

### Enrollment

Enrollment for all three studies was based on call census at the TMDS which was estimated prior to the INACT2-H and adjusted following each completed study^10,11^.

### Participant incentives and fees

Across all studies TMDS participants were asked to pay a fee of 500 Gourdes (5 USD) to contribute to the cost of delivery and medications. The fee was reduced or waived for those who were unable to pay in full. The funds were re-invested in the study and the research was not dependent on the fee. The fee was implemented to avoid incentivizing families to delay daytime care for free nighttime care and to avoid undercutting daytime providers who charge a fee.

### Implementation

The TMDS workflow, clinical procedures and TMDS guidelines for INACT2-H and INACT3-H are described previously^10,11^ and the ongoing INACT4-H protocol is included in appendix 2. The workflow (**Figure 1η)** steps are: i) A parent contacted the call center. ii) The provider gathered information about the child using a paper case report form (CRF) that included elements of decision support and generated a treatment plan that consisted of a disease severity classification (mild, moderate, or severe), disposition (hospital or household), recommended medications and/or fluids, and advice on follow-up care. iii) Severe cases were immediately referred to the hospital. iv) For non-severe cases, patients in a delivery zone received a medication delivery with or without an in-person exam. For INACT2-H, all cases had a paired exam. For INACT3-H and INACT4-H, only cases with higher clinical uncertainty (mainly moderate cases) received an in-person exam whereas the remainder received a medication delivery alone. v) Children residing outside a delivery zone, received a virtual exam and advice alone. vi) Patients requiring hospital or mandatory clinic follow-up received a 24-hour follow-up call. All patients received a 10-day call to report the child’s condition and provide feedback.

**Figure 1.**
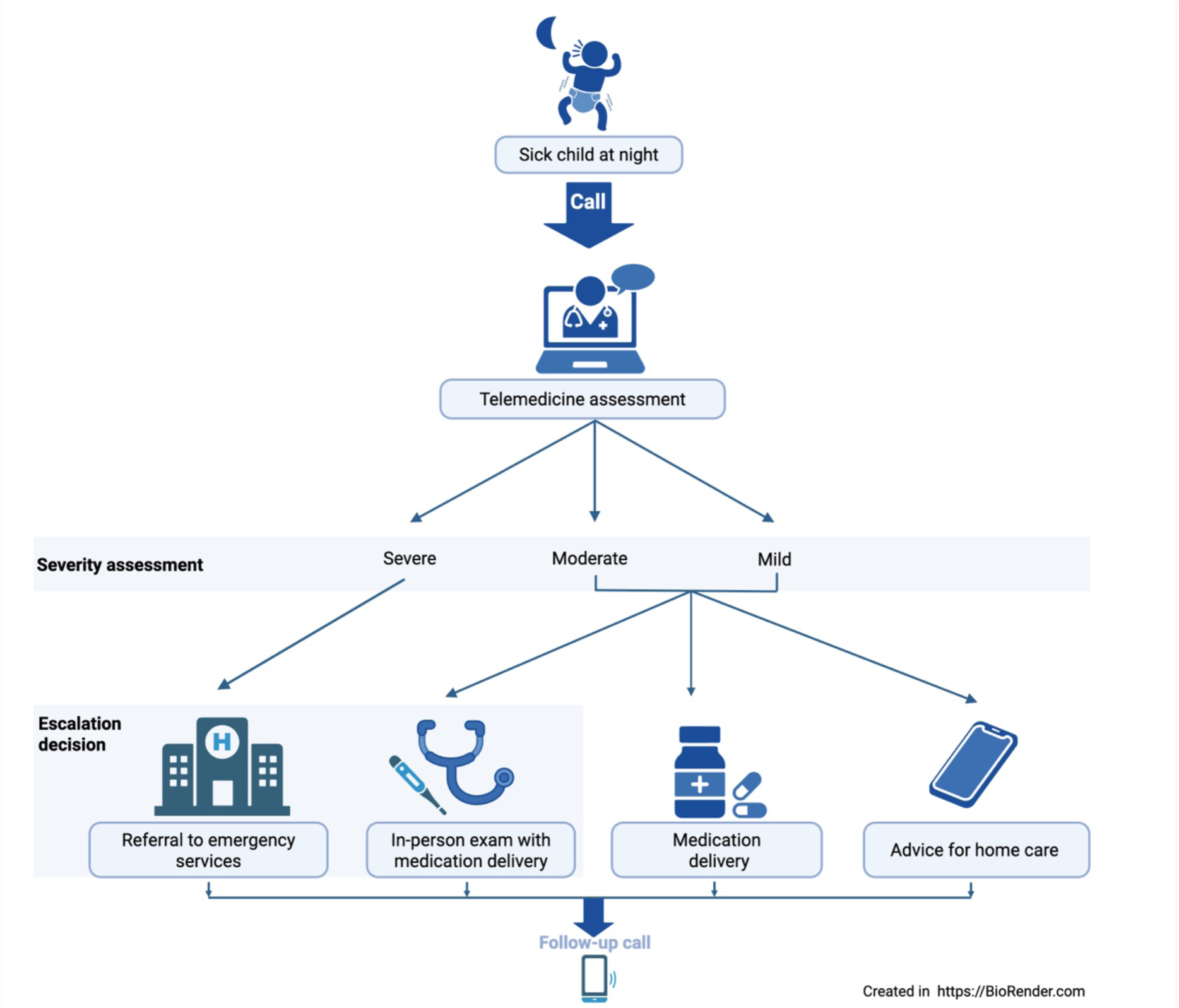
General workflow of the telemedicine and medication delivery service (TMDS). The parent of a child who is sick at night contacts the call center. A TMDS provider conducts a virtual exam with questions on clinical context. The provider references clinical guidelines and resources to make and assessment and create a treatment plan. The plan is based on the illness severity assessment which informs the decision to escalate or not escalate the disposition. Escalation includes hospital/ emergency service referral for severe cases, or cases with higher clinical uncertainty (mostly moderate) receive an in-person exam with medication delivery. Cases with low clinical uncertainty (mostly mild) receive medication delivery or advice for home care alone if a delivery is not indicated or feasible. Families outside the delivery zone receive anticipatory advice. All families are contacted by phone at 24-hours and/or 10-days for follow-up. In INACT2-H all non-severe cases residing within the delivery zone received an in-person household exam for research purposes and medication delivery (alone) was not part of the workflow.

### Outcome measures

#### Primary

The primary outcome measure in this secondary analysis was prediction of disease severity (mild vs not-mild) based on features collected via virtual exams at the call center. The primary metric was area under the receiver operating characteristic curve (AUC). Based on academic standards, a threshold of 0.80 or greater was considered ‘good’ performance^15^.

#### Secondary

The secondary outcome measure was prediction of the need for escalation of care (in-person exam at the household or hospital referral).

#### Exploratory

The relative contributions of individual features within the models were evaluated. Enumerations of cases that would be flagged as non-mild or needing escalation were estimated from the population based on a given probability generated by the models.

### Data curation

We combined data on participants with paired virtual and in-person exams from the INACT-2H, INACT-3H, and INACT-4H (through August 2023) studies^10,11^. The date cutoff was chosen because records to this point were complete at the time model derivation was initiated. Records with missing data for primary outcome variables (disease severity or disposition) were removed. For all studies, disposition during in-person exams, and for INACT2-H disposition during virtual exams, was determined manually based on the clinical guidelines. The severity of cases with missed ‘danger signs’ during either the virtual or in-person exams were manually reassigned to severe. ‘Danger signs’ are clinical indicators (e.g., severe dehydration) outlined in the clinical guidelines (appendix 3) that signify a case is severe.

Given that data were collected from multiple studies, there were discrepancies amongst the variables across studies. Moreover, within a study, some questions were only asked to certain patients based on branching logic. For example, INACT-2H did not ask whether the parent was concerned about the child’s weight and in INACT-3H and INACT-4H this question was only asked if the child was less than 60 months old. When appropriate, we used branching logic to fill in some of the missing values. For example, if the parent did not report diarrhea as a complaint, then the number of diarrheal episodes in the previous 24 hours was imputed as 0, rather than missing. However, some instances of missing data could not be resolved through logical imputation, e.g., questions that were not asked across all studies. In such cases, we left those values as missing. The XGBoost’s built-in handling procedure which learns the best way to route missing values during the tree-building process, eliminates the need for explicit imputation.

Given that virtual exams were conducted over the phone and relied on the parent correctly hearing and understanding questions, providers were able to document instances where they felt the information they were receiving might be unreliable by marking the response as “unsure” on the CRF. In these situations, the question was omitted when generating the treatment plan. We accounted for ‘unsure’ responses using the following methods: For binary variables such as ‘blood in stool’, we created an ordinal variable with levels “No”, “Yes and unsure” and “Yes”. For ordinal variables such as number of vomiting episodes in the last 24h, we created an “unsure” indicator variable as a separate feature in the model. For chief complaint, a categorical variable with more than two categories, we created dummy variables for each level with 0 for all representing a missing chief complaint.

### Model derivation and validation

We developed disease severity prediction (DSP) models for two binary outcomes: 1) Not-mild severity, the primary outcome, was defined as positive for patients classified as moderate or severe and 2) Escalation of care, defined as positive if a patient requires an in-person provider visit or referral to emergency services. We only considered variables that were collected during the virtual exam and those that were collected during the in-person exam (e.g., oxygen saturation) were excluded. We tested models that included the “unsure” indicator variables and ordinal variables with “unsure” as a level, as well as models that excluded these variables where ‘yes and unsure’ and ‘no and unsure’ were treated as ‘yes’ and ‘no’, respectively.

We used the XGBoost algorithm, an ensemble approach that uses gradient boosted decision-trees to derive predictive models for estimating the probability that a TMDS case would be ‘not-mild’ or require escalation. We used five-fold cross-validation to tune hyperparameters: max depth, η (learning rate), subsample, column sample by tree, lambda, and number of rounds (trees). We considered 100 rounds (or trees) in each model fit with early stopping if not improved over 10 rounds. Area under the receiver operating characteristic curve (AUROC or AUC) was used to assess performance for both cross-validation, model fitting, and importance metrics. Given the small sample size of our data and the number of features considered (35), we opted to restrict our analysis to 5-fold cross-validation rather than a separate validation set.

While the model is trained to maximize AUC, we also report the area under the precision-recall curve (PRAUC) and the calibration of the model which assesses how well the predicted risk aligns with the observed risk. We calculated the importance of each predictor using built-in XGBoost output which calculates the fractional contribution of each predictor to the model performance (log-loss) based on the total gain in performance when its decision tree splits were utilized. A higher percentage indicates a more important predictor. Finally, Shapley Additive Explanations (SHAP) were calculated, a value that expresses the additive contribution of each feature to the predicted value and is used to explain the direction and magnitude of a predictor on the outcome dependent on its value.

## RESULTS

### Participant characteristics

A total of 2225 cases were enrolled between the three studies; the median age was 24 months and 47% were female. Given differences in study designs, there were 683 cases with paired virtual and in-person exams available for analysis; the median age was 15 months and 47% were female. Additional participant characteristics are shown (**Table 1**). Features that may have differed between the virtual (call center) and in-person (household) settings were enumerated.

**Table 1.**
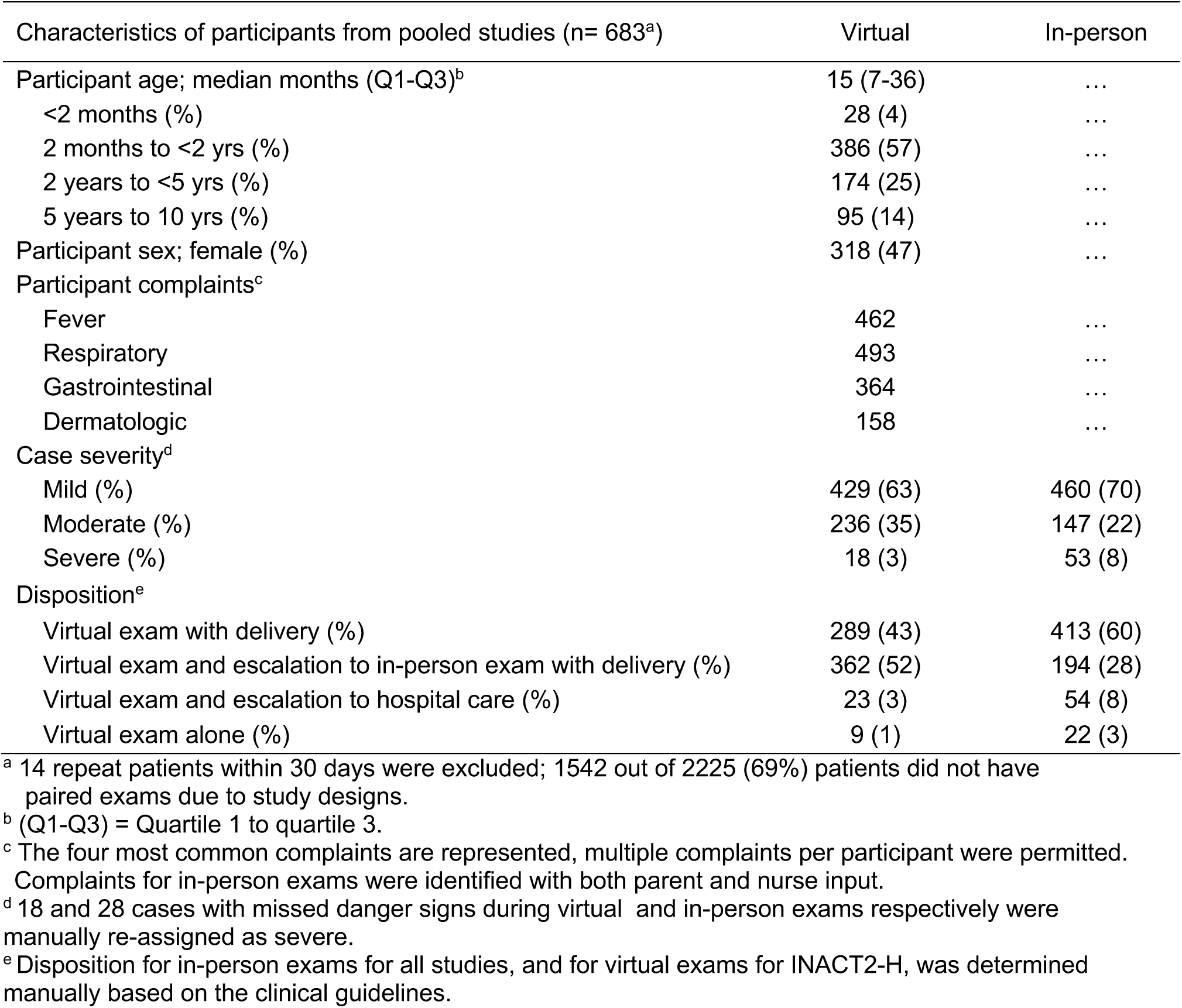
Characteristics of participants with paired virtual and in-person exams.

### Primary outcome

The models were computed both by accommodating for and not accommodating for providers’ perception of the reliability of responses, or the ‘unsure’ designation. When the ‘unsure’ designation was not considered, the model to predict the detection of a ‘not-mild’ status had an AUC of 0.82 (95% CI 0.78-0.86) after 5-fold cross validation; calibration slope and intercept were 1.09 (95% CI: 0.91-1.26) and 0.16 (95% CI: -0.03 – 0.35), respectively **(ηFigure 2η)**. When the ‘unsure’ designation was considered, the model to predict the detection of a ‘not-mild’ status had an AUC of 0.82 (95% CI 0.78-0.85) after 5-fold cross validation; calibration slope and intercept were 0.91 (95% CI: 0.76 -1.06) and 0.2 (95% CI: 0 – 0.4), respectively (**appendix 1 p 2**).

**Figure 2.**
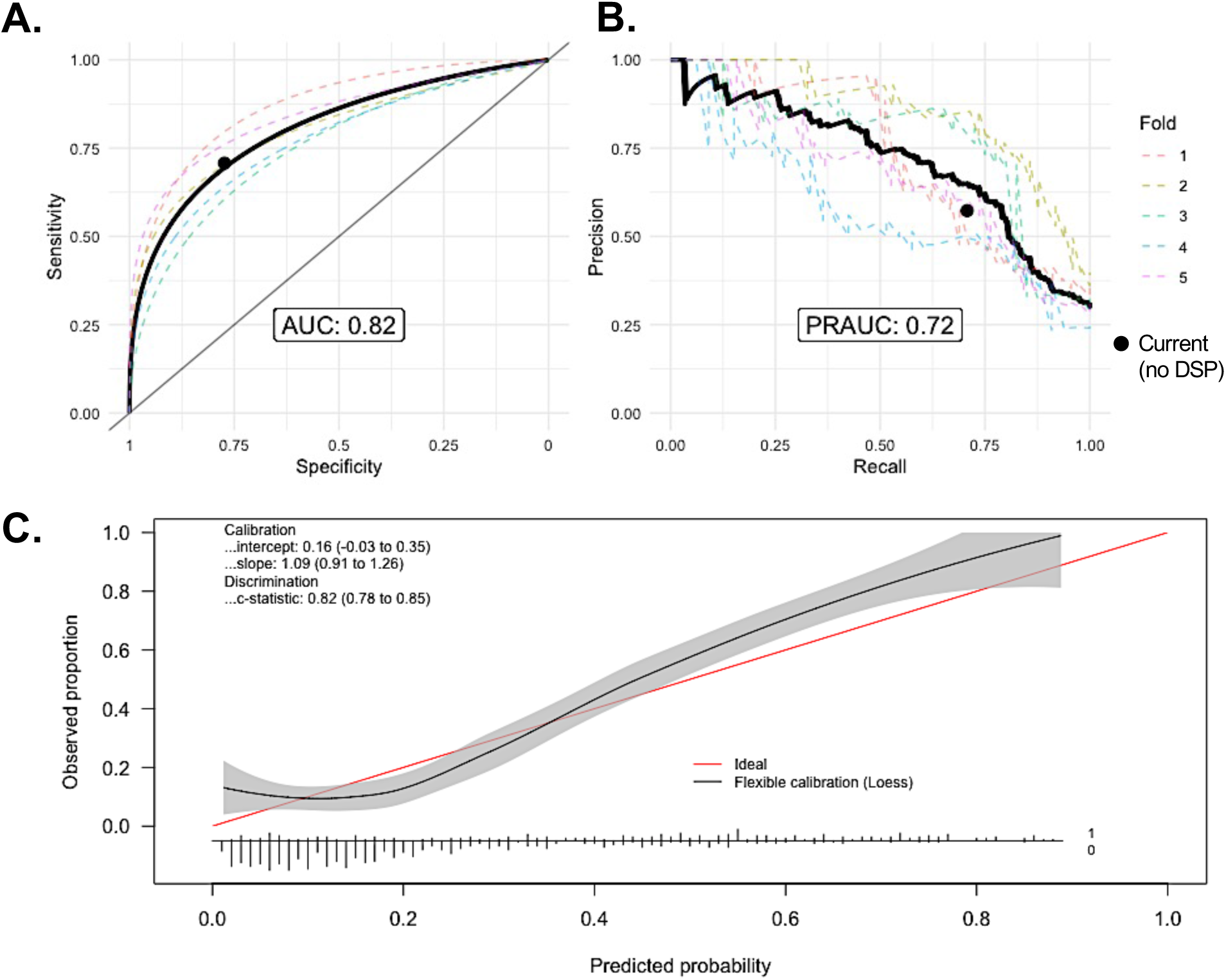
Prediction of disease severity. Receiver operating characteristic curve. (B) Precision-recall curve. (C) Calibration curve for the disease severity predictive model. In plots A and B, the individual dashed lines represent testing performance of each of the 5-folds and the thick black line represents the combined folds curve. The calibration plot (C) is only based on the combined predictions for all five folds and shows a histogram of prediction density by true outcome (0 – Mild, 1 – Severe) at the bottom. Black circle represents current sensitivity and specificity of provider using the clinical guidelines without the DSP.

### Secondary outcome

For prediction of escalation of care that does not consider the ‘unsure’ designation, the model had an AUC of 0.78 (95% CI 0.74-0.81) after 5-fold cross validation; the calibration slope and intercept were 0.81 (95% CI: 0.66-0.96) and 0.08 (95%CI: -0.11-0.26), respectively **(ηFigure 3η)**. When considering the ‘unsure’ designation, the model had an AUC of 0.78 (95% CI 0.73-0.81) after 5-fold cross validation; calibration slope and intercept were 0.47 (95% CI:0.39 -0.56) and 0.11 (95%CI: -0.11 – 0.33), respectively (**appendix 1 p3**).

**Figure 3.**
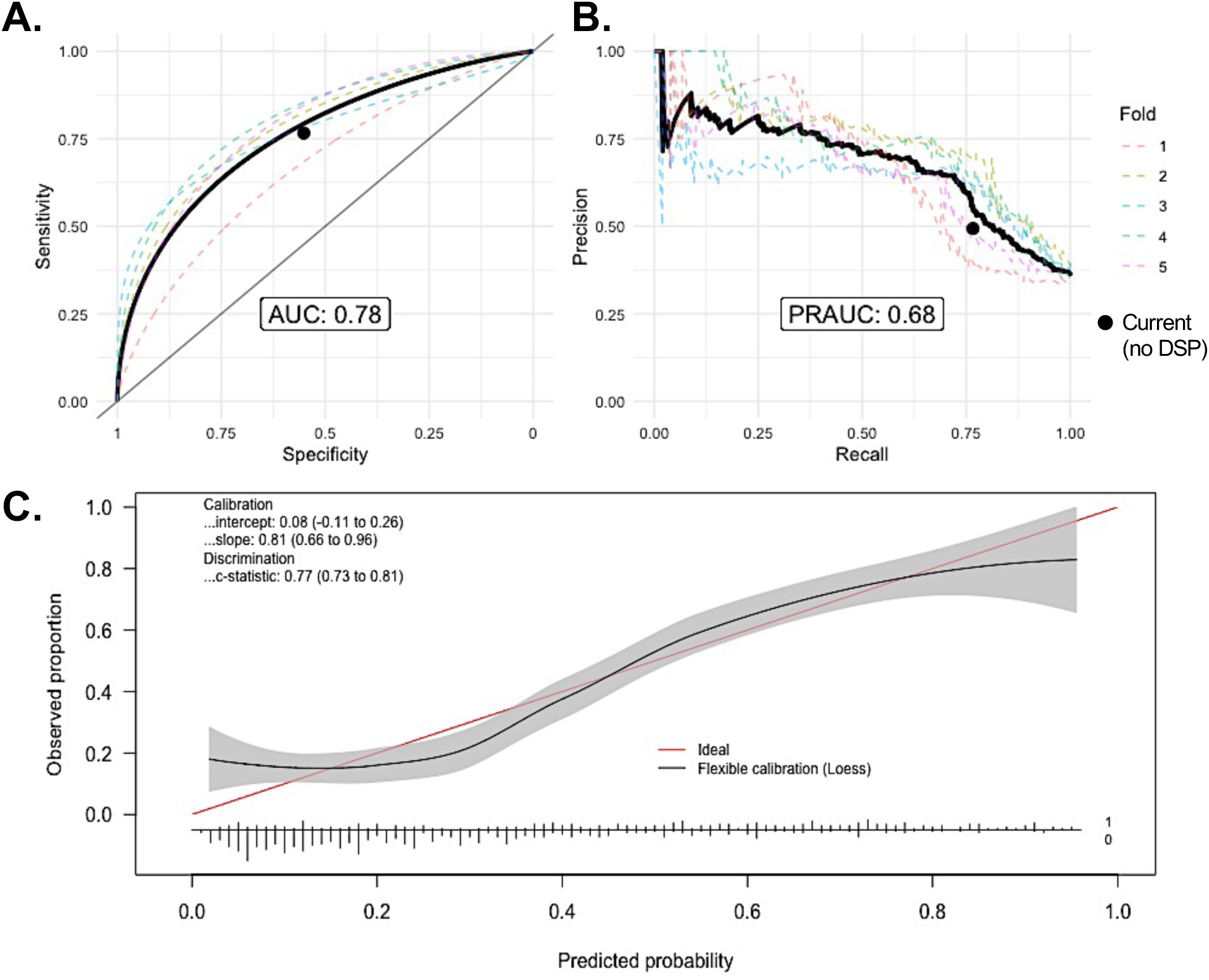
Prediction of need for escalation. Receiver operating characteristic curve. (B) Precision-recall curve. (C) Calibration curve for the escalation of care predictive model. In plots A and B, the individual dashed lines represent testing performance of each of the 5-folds and the thick black line represents the combined folds curve. The calibration plot (C) is only based on the combined predictions for all five folds and shows a histogram of prediction density by true outcome (0 – No Escalation, 1 – Escalation) at the bottom. Black circle represents current sensitivity and specificity of provider using the clinical guidelines without the DSP.

### Exploratory analyses

SHAP values were used to assess the additive contribution of each feature to the predicted value of a given model; these values and associated plots help explain the direction and magnitude of a predictor on the outcome. We show the top ten most important variables and SHAP values for each model that did not consider the ‘unsure’ designation (**appendix 1 p4-5)**.

Estimated case counts flagged ‘up’ for additional consideration by the DSP models are shown (**Table 2**). When applying the severity prediction model at a balanced sensitivity of 75% and specificity of 80%, 11% of cases would be flagged ‘up’ for additional consideration, of which 38% would be ‘true’ positives. Similarly, when applying the escalation prediction model at a balanced sensitivity of 75% and specificity of 71%, 11% of cases would be flagged for escalation, of which 36% percent would be ‘true’ based on in-person. While outside the intended implementation goals for safety, larger effects were observed to flag cases ‘down’ which may have benefits for cost savings.

**Table 2.**
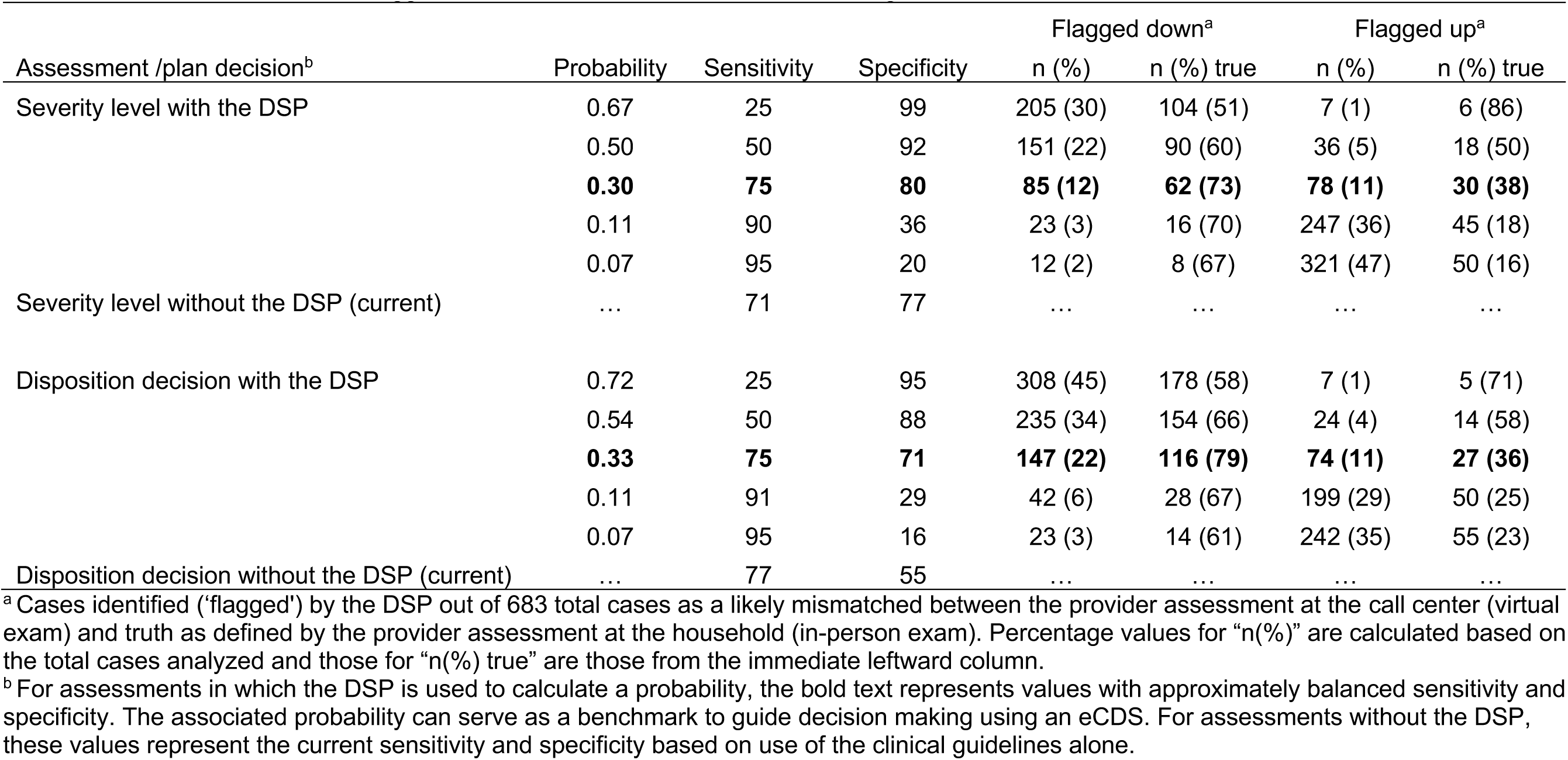
Estimated case counts flagged for additional clinical consideration during virtual exams at a TMDS call center.

| Assessment /plan decision<sup>b</sup> | Probability | Sensitivity | Specificity | Flagged down<sup>a</sup> |  | Flagged up<sup>a</sup> |  |
| --- | --- | --- | --- | --- | --- | --- | --- |
|  |  |  |  | n (%) | n (%) true | n (%) | n (%) true |
| Severity level with the DSP | 0.67 | 25 | 99 | 205 (30) | 104 (51) | 7 (1) | 6 (86) |
|  | 0.50 | 50 | 92 | 151 (22) | 90 (60) | 36 (5) | 18 (50) |
|  | **0.30** | **75** | **80** | **85 (12)** | **62 (73)** | **78 (11)** | **30 (38)** |
|  | 0.11 | 90 | 36 | 23 (3) | 16 (70) | 247 (36) | 45 (18) |
|  | 0.07 | 95 | 20 | 12 (2) | 8 (67) | 321 (47) | 50 (16) |
| Severity level without the DSP (current) | ... | 71 | 77 | ... | ... | ... | ... |
| Disposition decision with the DSP | 0.72 | 25 | 95 | 308 (45) | 178 (58) | 7 (1) | 5 (71) |
|  | 0.54 | 50 | 88 | 235 (34) | 154 (66) | 24 (4) | 14 (58) |
|  | **0.33** | **75** | **71** | **147 (22)** | **116 (79)** | **74 (11)** | **27 (36)** |
|  | 0.11 | 91 | 29 | 42 (6) | 28 (67) | 199 (29) | 50 (25) |
|  | 0.07 | 95 | 16 | 23 (3) | 14 (61) | 242 (35) | 55 (23) |
| Disposition decision without the DSP (current) | ... | 77 | 55 | ... | ... | ... | ... |

<sup>a</sup> Cases identified ('flagged') by the DSP out of 683 total cases as a likely mismatched between the provider assessment at the call center (virtual exam) and truth as defined by the provider assessment at the household (in-person exam). Percentage values for "n(%)" are calculated based on the total cases analyzed and those for "n(%) true" are those from the immediate leftward column.
<sup>b</sup> For assessments in which the DSP is used to calculate a probability, the bold text represents values with approximately balanced sensitivity and specificity. The associated probability can serve as a benchmark to guide decision making using an eCDS. For assessments without the DSP, these values represent the current sensitivity and specificity based on use of the clinical guidelines alone.

## DISCUSSION

Historically, accurately identifying the ‘sick’ patient early required exceptional bedside assessment skills combined with an expansive fund of knowledge. How one integrates technology to improve outcomes and efficiency in a cost-effective manner is today’s challenge. We present a use case specific to telemedicine in resource-limited settings. Our objective was to develop models that flag potentially ‘sick’ patients during call center encounters for additional consideration that might otherwise be missed. To address this challenge, we developed a study design that utilized data from paired virtual and in-person exams at a TMDS in Haiti. Using machine learning analytic techniques, we derived and validated a model that predicted, and therefore discriminated, patients with moderate or severe disease as well as a model that predicts if a case needs escalation. Model performance was assessed using the AUC metric, with results at or above commonly accepted thresholds for inclusion in clinical practice. The models require external validation prior to integration and evaluation in clinical trials. Once validated, the models may enable a critical safety check for experienced providers and a method to digitally convey expertise to new providers.

These findings can be interpreted at three levels: the methodologic proof of concept, the analytic outcomes, and insights on human-computer interaction. The formative phase of this research began with an evaluation of pediatric telemedicine guidelines to understand to what degree the process of identifying the ‘sick’ child was evidence-based in a virtual exam environment. There was a paucity of literature in which guidelines and approaches used gestalt borrowed from in-person exam experiences to navigate the telemedicine environment. In situations with limited connectivity and resources, there was a need for a new strategy. Therefore, in the initial INACT study we developed an approach to conduct paired virtual and in-person exams on all cases within the delivery zone that did not need immediate hospital referral. We hypothesized that we could train models using the virtual exam data as predictors and the in-person exam data as ‘truth’. While developed in a low-resource setting, we challenge teams with similar interests to test this approach in high-resource settings.

The analytic outcomes are difficult to interpret because there is no clear threshold at which a model is ‘proven’ accurate and meaningful without incurring unintended consequences. An AUC threshold of 0.80 or above is generally considered a reasonable metric to advance to an external validation phase with possible subsequent deployment. However, initial design work during model derivation exposed the need for two models. The WHO IMCI guideline is based on a ‘stoplight’ system of green (mild), yellow (moderate) and red (severe). In high-risk triage scenarios involving pediatric patients in a virtual telemedicine environment, an accurate severity assessment is critical. This was the primary objective and outcome of our study. However, the first actionable decision following a virtual assessment is deciding whether the care plan requires escalation through a hospital referral or in-person exam at the household. Or, whether medication delivery alone with anticipatory guidance is sufficient. This insight is essential to ensuring clinical safety in telemedicine as it scales. We explored how probability thresholds for when to take action could be set by enumerating rates of flagged cases at varying sensitivities and specificities. The next step is to simulate and model the effects of threshold settings, comparing high sensitivity, balanced sensitivity and specificity, and high specificity on clinical safety and monetary costs.

Human-computer interactions within healthcare are complex and evolving. There is an unproductive conversation that AI-enabled eCDS systems will replace providers. We chose a different approach that recognized the value of both the provider (biologic intelligence) and the computer (artificial intelligence). Using human centered design, we developed the TMDS case report form to enable providers to both record responses (reporter phase) as well as mark those responses that the provider considers to be unreliable (synthesis phase). This latter step captures a feature of ‘biologic intelligence’ that is frequently relegated. The clinical guideline follows a traditional decision tree format, in which variables marked as potentially unreliable were treated as negative responses. This approach has inherent limitations that the models can potentially account for. Accordingly, the models were derived with and without incorporating these data. The results were mixed and did not show a clear advantage, likely due to limited sample sizes at both the patient and provider levels. Nevertheless, we consider this an important open question in the field of human-computer interaction that warrants further investigation.

These findings should be viewed within the context of the study limitations. The first limitation relates to a generally small sample size. To address this limitation several steps were taken that included selecting a subset of predictive variables for model derivation. This may have created a bias in the models by unintentionally excluding factors with meaningful effects. Second, the models were derived using datasets pooled data from three different studies to increase sample size (INACT2-H, INACT3-H, INACT4-H), however the study population may have changed over the four-year period. Additionally, differences in study designs between INACT2-H and INACT3/4-H led to some variation in the types of cases that received paired exams. In INACT2-H, all non-severe cases had paired exams, whereas in INACT3/4-H, only cases with higher clinical uncertainty, primarily moderate cases, had paired exams. Although the majority of the CRF was consistent across studies, variations of certain variables may have affected model development. Third, the training dataset lacked a balanced distribution of mild, moderate, and severe cases. This imbalance is partly attributable to the study workflow which required hospital referral for severe cases identified during virtual exams. As a result, few severe cases were encountered during in-person exams. Our derivation methods were selected to account for this limitation. Forth, the decision tree embedded in the clinical guideline uses a singular triage strategy (green, yellow, red) based on WHO methods. While simplified, triage is based on both physiologic and logistical considerations. The models reflect this feature and disaggregating the physiologic and logistical elements was not possible with the approach taken. This presents a contextual limitation that may constrain the model’s applicability to settings with different operational or triage workflows.

## CONCLUSION

This study presents an important proof-of-concept for how to develop evidence-based pediatric telemedicine guidelines enhanced with predictive models that prioritize safety for vulnerable populations, especially children in resource limited settings. The models require external validation prior to eCDS integration. Once validated, the models are intended to provide a critical safety check for experienced providers and digitally convey expertise to new providers.

## Data Availability

All data used in the present study are available upon reasonable request to the authors.

## Abbreviations and acronyms

TMDS: Telemedicine and medication delivery service
WHO: World Health Organization
IMCI: Integrated Management for Childhood Illness
CRF: Case report form
INACT: Improving Nighttime Access to Care and Treatment
IQR: Interquartile range

## Contributions

Conceptualization- BJB, MBK, EJN

Data curation- BJB, MBK, YC, KF

Formal analysis- BJB, MBK

Funding acquisition- EJN

Investigation- MBK, YC, LE, JRB, CB

Methodology- BJB, MBK, EJN

Project administration- MBK, EJN, VMBR

Supervision- MBK, YC

Visualization- BJB, MBK, EJN

Writing – original draft- BJB, MBK, EJN

Writing – review & editing- BJB, MBK, EJN, LE, CB, JRB, VMBR, KF, YC

## Acknowledgments

We would like to thank both the study participants and the TMDS staff for their contributions to the datasets analyzed in this manuscript. We are grateful to our University of Florida administrator team R Autrey, K Berquist, B Johnson, T Linn, and N Rushing. We are thankful for the academic support provided by G. Morris at the Emerging Pathogens Institute, and R. Savani Chair of the Department of Pediatrics, as well as our colleague T. Becker at the University of Florida for guidance and clinical guideline reviews. J. Friesen at Trek Medics International has provided important guidance and access to logistical software that is used in the operation aspects of MotoMeds. Finally, we acknowledge that our partnership with the Ministry of Public Health and Population (Ministère de la Santé Publique et de la Population - MSPP) in Haiti is invaluable to this work.

## Data Sharing

De-identified individual participant data and the data key that underlie the results are available in appendix 4.

## Supplementary Materials

**Appendix 1**.

**Figure S1.**
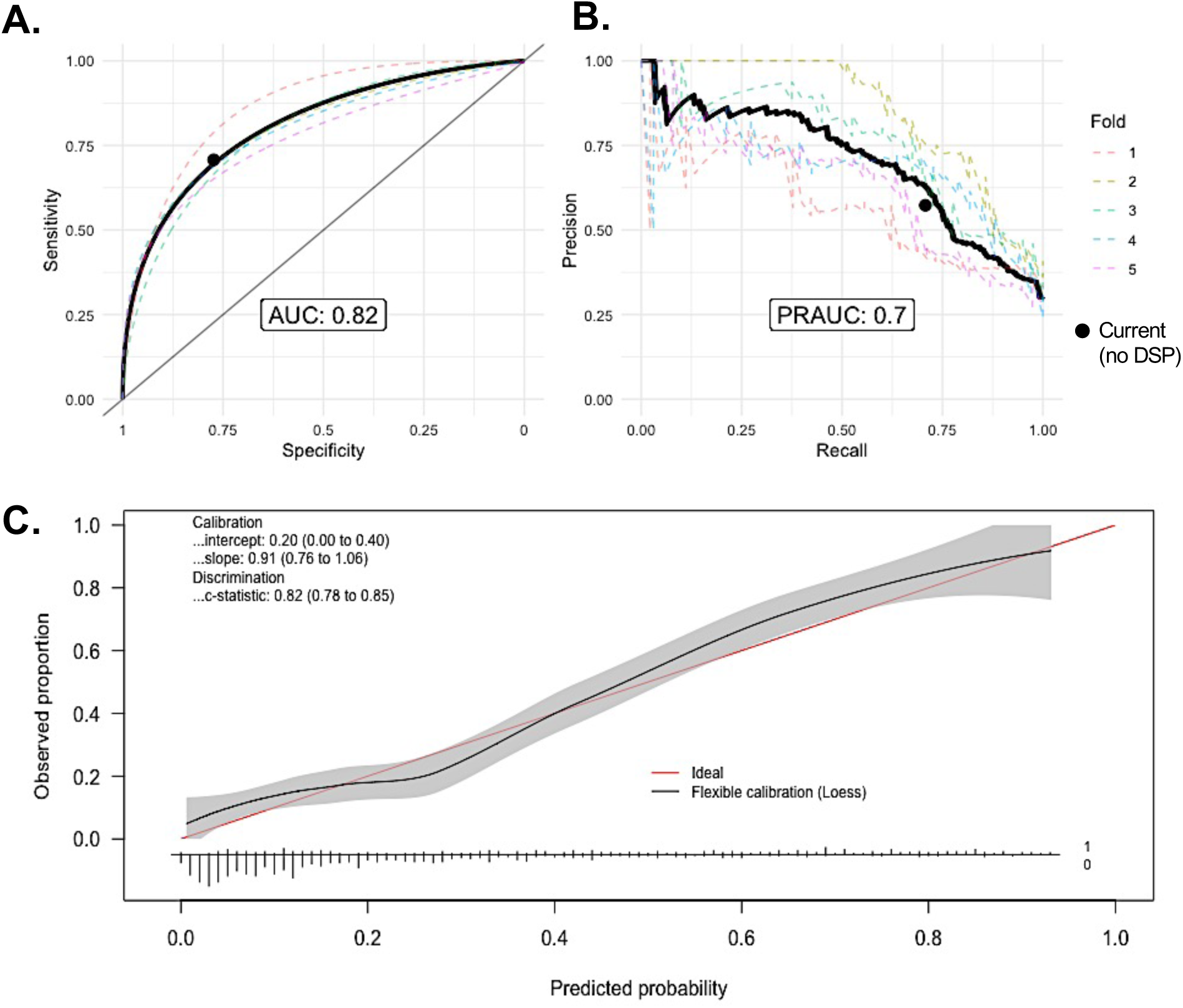
Prediction of disease severity when considering ‘unsure designation’. (A) Receiver operating characteristic curve. (B) Precision-recall curve. (C) Calibration curve for the disease severity predictive model. In plots A and B, the individual dashed lines represent testing performance of each of the 5-folds and the thick black line represents the combined folds curve. The calibration plot (C) is only based on the combined predictions for all five folds and shows a histogram of prediction density by true outcome (0 – Mild, 1 – Severe) at the bottom. Black circles represent current sensitivity and specificity of provider using the clinical guidelines without the DSP.

**Figure S2.**
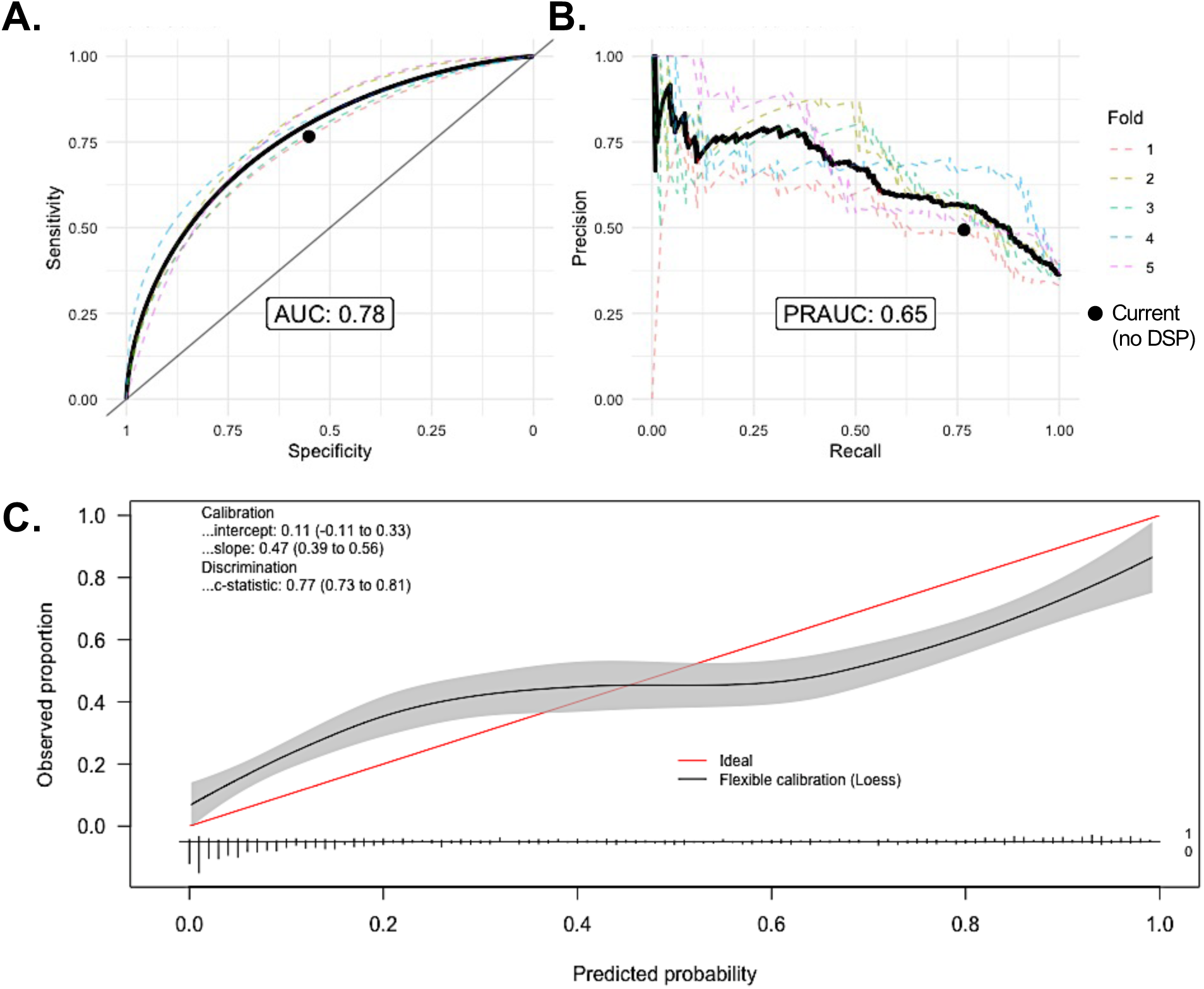
Prediction of need for escalation when considering ‘unsure designation. Receiver operating characteristic curve. (B) Precision-recall curve. (C) Calibration curve for the escalation of care predictive model. In plots A and B, the individual dashed lines represent testing performance of each of the 5-folds and the thick black line represents the combined folds curve. The calibration plot (C) is only based on the combined predictions for all five folds and shows a histogram of prediction density by true outcome (0 – No Escalation, 1 – Escalation) at the bottom. Black circles represent current sensitivity and specificity of provider using the clinical guidelines without the DSP.

**Figure S3.**
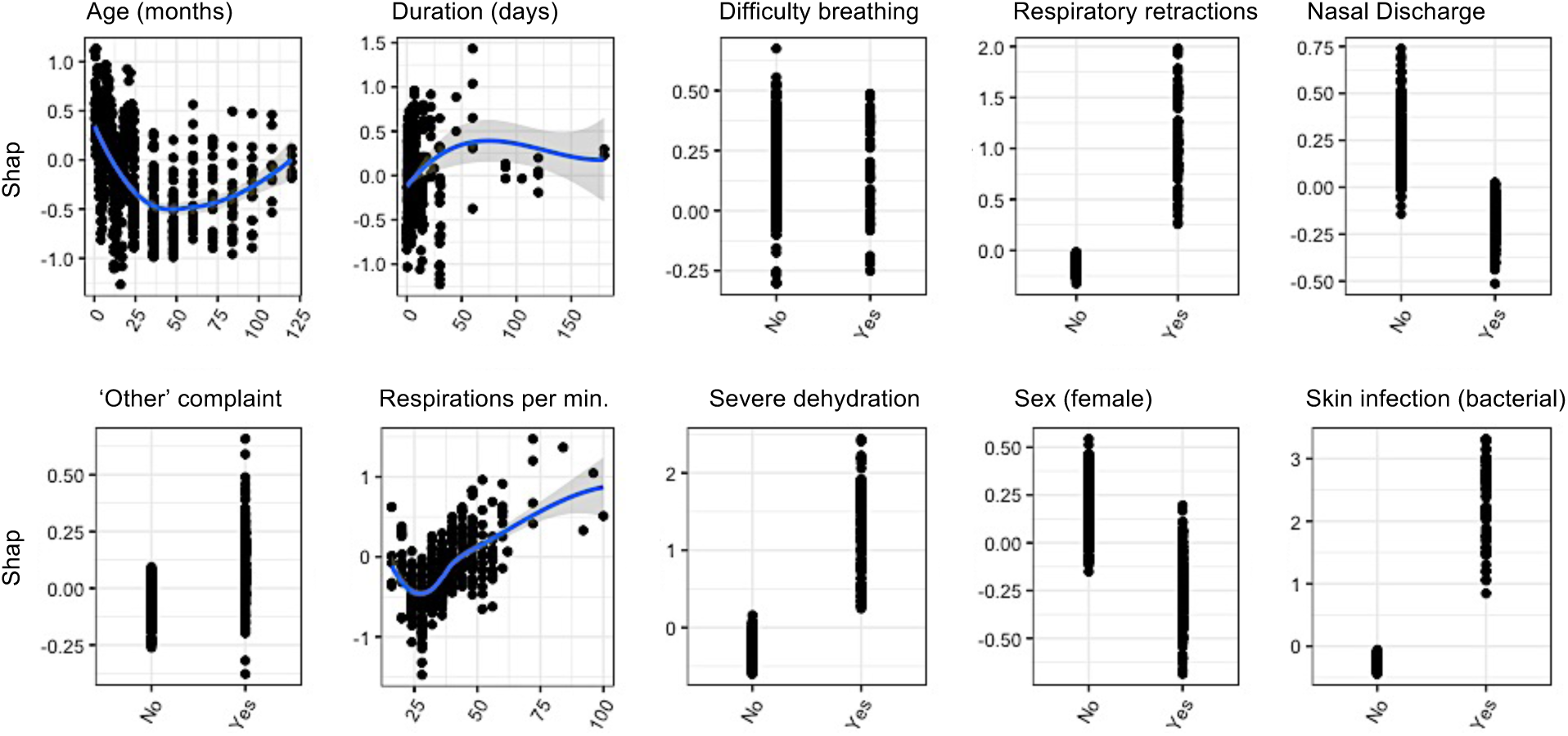
Contribution of individual features to the final model prediction for severity level. The ten most impactful individual predictors. Each feature’s contribution was assessed using Shapley Additive Explanations (SHAP) values which explain the direction and magnitude of a predictor’s effect based on its value. The plots are ordered by their relative importance from left to right and top to bottom, measured as the fractional contribution of the respective feature to the model fit (log-loss) based on the total gain of the feature’s splits. For continuous variables, the 95% confidence intervals are shown.

**Figure S4.**
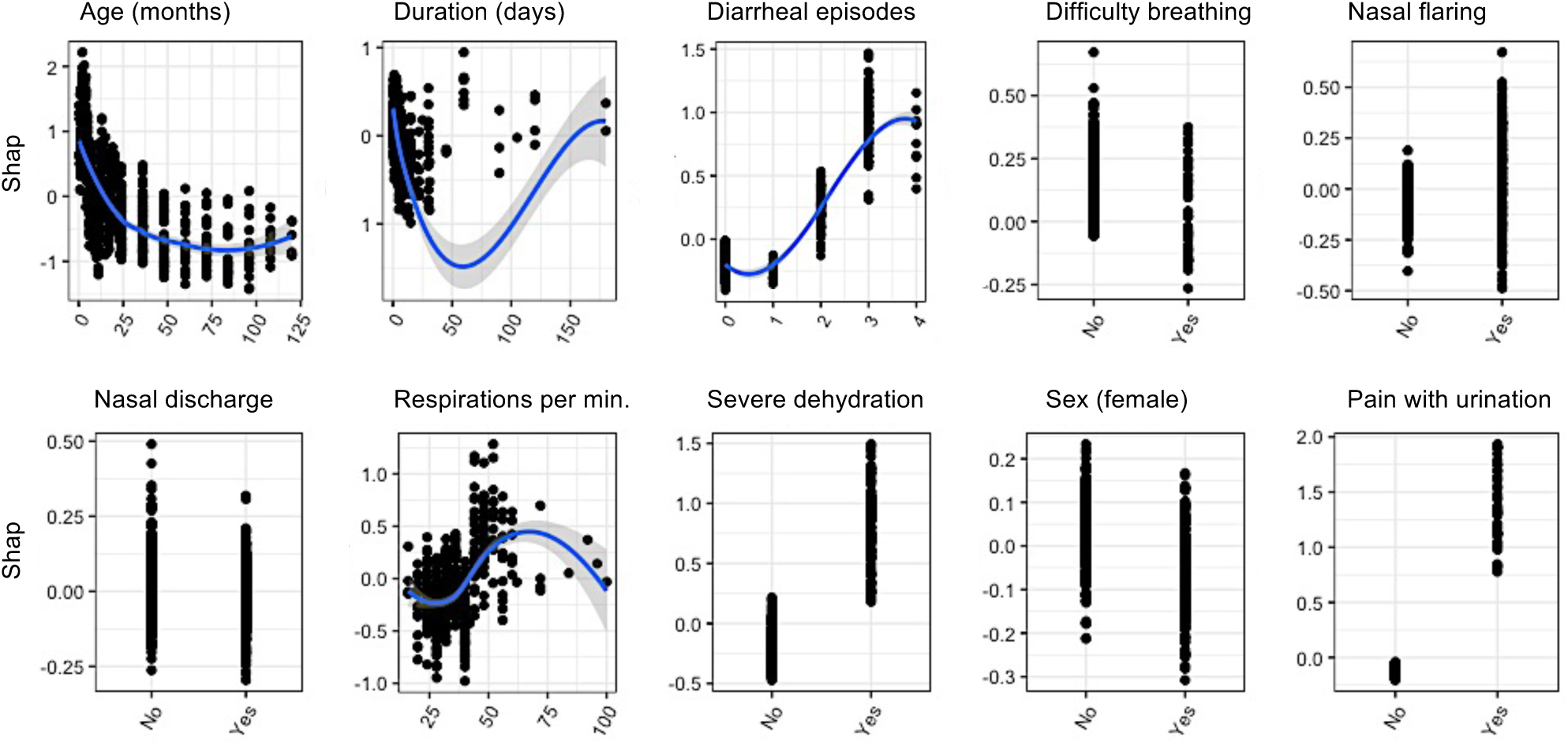
Contribution of individual features to the final model prediction for escalation. The ten most impactful individual predictors. Each feature’s contribution was assessed using Shapley Additive Explanations (SHAP) values which explain the direction and magnitude of a predictor’s effect based on its value. The plots are ordered by their relative importance from left to right and top to bottom, measured as the fractional contribution of the respective feature to the model fit (log-loss) based on the total gain of the feature’s splits. For continuous variables, the 95% confidence intervals are shown.

**Appendix 2.** INACT Study Protocol (INACT4H version)*

**Appendix 3.** INACT Clinical Guidelines (INACT4H version)*

**Appendix 4.** De-identified dataset and data key*

**Appendix 5.** Statistical code*

*AVAILABLE UPON REQUEST AND PROVIDED AT PUBLICATION IN JOURNAL

